# Face Coverings and Respiratory Tract Droplet Dispersion

**DOI:** 10.1101/2020.08.11.20145086

**Authors:** Lucia Bandiera, Geethanjali Pavar, Gabriele Pisetta, Shuji Otomo, Enzo Mangano, Jonathan R. Seckl, Paul Digard, Emanuela Molinari, Filippo Menolascina, Ignazio Maria Viola

## Abstract

Respiratory droplets are the primary transmission route for SARS-CoV-2. Evidence suggests that virus transmission can be reduced by face coverings, but robust evidence for how mask usage might affect safe distancing parameters is lacking. Accordingly, we investigate the effectiveness of surgical masks and single-layer cotton masks on mitigating dispersion of large respiratory droplets (i.e. non aerosol). We tested a manikin ejecting fluorescent droplets and human volunteers in speaking and coughing conditions. We quantified the number of droplets in flight using laser sheet illumination and UV-light for those that had landed at table height at up to 2m. For human volunteers, expiratory droplets were caught on a microscope slide 5cm from the mouth. Whether manikin or human, wearing a face covering decreased the number of projected droplets by >1000-fold. We estimated that a person standing 2m from someone coughing without a mask is exposed to over 1000 times more respiratory droplets than from someone standing 5 cm away wearing a basic single layer mask. Our results indicate that face coverings show consistent efficacy at blocking respiratory droplets. If aerosol transmission is later determined to be a significant driver of infection, then our findings may overestimate the effectiveness of face coverings.

## 1. Introduction

SARS-CoV-2 is primarily transmitted from virus-laden fluid droplets ejected from the mouth of an infected carrier. These droplets are either inhaled by a recipient, deposited on the recipient’s mouth or conjunctiva, or deposited on a surface (thereby generating a fomite) and then mechanically transmitted through physical contact [1]. The relative importance of these mechanisms is not proven, but evidence indicate that the predominant route is through droplet inhalation or direct deposition on the recipient’s mouth, nose or conjunctiva [2-4].

Recent reviews [5-7] suggest that face coverings are effective in decreasing the risk of infection. This has been inferred through epidemiological studies [8], tests with animals [9], and physical tests. For example, Viola et al. [10] showed that face coverings decreased the front throughflow of the aerosol jet by about an order of magnitude.

Comparatively, there is weaker evidence that face coverings mitigate dispersion of large respiratory droplets, although these seem to be the main route of virus transmission [2-4]. Palacios et al. [11] sprayed bacterial-suspension droplets with a diameter from 20 μm to 900 μm through different household textiles and showed that this could mitigate large droplet dispersal. Yet, to allow an evidence-based assessment of what the appropriate social distance is when masks are worn, face fitted masks must be tested with realistic respiratory airflow jets and with human volunteers.

Recently, Anfinrud et al. [12] demonstrated that large respiratory droplets ejected by a person speaking can be visualised by laser sheet illumination. Successively Fisher et al. [13] used this technique to compare the effectiveness of different face masks in filtering respiratory droplets. A person spoke in front of a hole in an enclosed box, where droplets passing through a laser sheet were imaged and counted for different face coverings. These experiments demonstrated that face coverings can be effective in mitigating the dispersion of droplets ejected by a person speaking. On the other hand, these studies did not provide information on the distance travelled by the droplets, and thus to how face coverings can be used to review social distancing guidelines.

We adopted a similar technique to quantify the in-flight droplets ejected by an anatomically realistic manikin, with and without face covering, at several distances up to 2 m from the source. We consider both speaking and coughing conditions. Furthermore, we complemented these measurements with two other independent measuring techniques, UV light imaging and microscopy, which resulted in consistent results. Finally, we measured with microscopy the droplets ejected by six individuals and the results corroborated those of the manikin.

## 2. Methods

### 2.1 SPEECH AND COUGH SIMULATOR

The same simulator used by Viola et al. [10] was used. Air flow was generated using an air compressor capable of delivering up to 180 litre/min, while a 100 μM fluorescein solution in water for droplet generation was supplied by a TCS M400S micropump with a variable output range of 0-2.7 litre/min. Liquid and air flows were connected to a purpose-built droplet generation system fitted inside an anatomically realistic, adult, medical simulation manikin’s mouth (Resusci Anne QCPR) that creates controllable droplet sizes of up to about 1 mm. Air/liquid flows were ejected from a 2 cm diameter circular “tracheal” opening with a flow rate and velocity comparable to those of a person either speaking [14] (1 m s^−1^) or coughing [15] (10 m s^−1^). Masks were either surgical or single layer woven cotton (see Supplementary Material). Examination of samples by phase contrast microscopy showed that the cotton masks had a close weave with gaps of around 50 μm, while the multiple layers of less densely woven material in the surgical mask left a broader spectrum of pore sizes (Figure S1).

### 2.2 LASER IMAGING OF DROPLETS IN FLIGHT

We projected a thin laser sheet along the vertical plane through the mouth of the manikin and used a photographic camera to capture the light scattered by droplets passing within this plane (Supplementary Movie 1). A 2.5 W diode-pumped continuous wave laser (532 nm) running at 40% of maximum power illuminated a plane perpendicular to the floor along the air flow jet axis. An 8-bit CCD camera with a resolution of 2056 × 2060 pixels with a Nikon 50 mm f/2 lens was used to image a physical plane of 137.5 × 137.8 mm, with a resolution of 67 μm/pixel. A 60 ms exposure time was used to count particles under all conditions except coughing without a mask, where 30 ms was used. A total of 100 images from six replicate experiments were used for particle counting analysis. The light scattered by fluorescein made droplets appear larger than their actual size. Using a shadow imaging technique, we verified that the laser imaging visualised all droplets large enough to fall ballistically (see Supplementary Material).

### 2.3 UV-LIGHT IMAGING

Paper sheets were placed on a table 0.426 m below the manikin to cover areas of 0.84 m × 0.6 m for speaking and 2.1 m × 0.3 m for coughing. Speaking and coughing tests without a face covering lasted two and one minute, respectively, while durations of 20 and 10 minutes were used when the manikin was wearing a handmade or surgical mask. Each condition was tested with at least six replicates. After each test, paper samples were placed on a Safe Imager 2.0 Blue-Light Transilluminator (InvitrogenTM G6600) to visualise deposited droplets. Images were acquired using an iPhone 7 camera (f/1.8, 4032 × 3024 pixels) with a resolution of 65 μm/pixel. Images were imported into Inkscape to reconstruct the continuous sample and to equalise the pictures across the whole sample. Upon binarisation of the reconstructed image with a manually defined threshold, droplets were counted in each grid element using Fiji’s edge-detection algorithm.

### 2.4 MICROSCOPY

Samples caught on glass microscope slides were imaged with a CFI Plan Fluor 20X objective mounted on a Nikon Eclipse Ti inverted microscope, equipped with an iXon Ultra 888 EMCCD camera, resulting in a resolution upon magnification of 0.65 μm/pixel. For each sample, 210 fields of view, covering an area of 0.85 cm^2^ in total, were acquired in the FITC channel (Gain 70, Exposure time 100 ms) using NIS-Elements. Samples collected in tests involving human subjects were imaged with an Andor Zyla sCMOS camera and the same objective and microscope as above, resulting in a resolution upon magnification of 0.33 μm/pixel. We acquired 153 fields of view, corresponding to 0.62 cm^2^. All images were downscaled by a factor of five prior to their analysis in scikit-image, Python. Droplets in each sample were counted as the markers of the watershed algorithm, which allows segmentation of touching objects in an image. A Gaussian smoothing filter was applied (standard deviation of five) to reduce pixel noise before a thresholding procedure using a manually selected value. Following morphological opening with a Boolean kernel of size seven, droplet centres were identified as the local maxima of the distance transform of the image, with the stipulation that centres should be more than 15 pixels from each other.

### 2.5 TESTS WITH HUMAN VOLUNTEERS

Six volunteers performed two rounds of coughing (1 min/each) and speaking (3 min/each) tasks, with and without a surgical mask. Speech tasks were performed reading a provided sample text, to ensure results were not biased by the personal choice of words. On the two days of the trial experiment, the order in which the tasks were performed was reversed. This research protocol was approved by the University of Edinburgh’s Human Research Ethical Review Committee and all human participants gave written informed consent.

### 2.6 STATISTICAL ANALYSIS

The mean and the standard error of the mean (SEM) of the data are presented. Due to inhomogeneity of variance, results were assessed by Bonferroni-corrected Kolmogorov-Smirnov statistical tests applied to pairwise combinations of no-mask/handmade mask, no-mask/surgical mask, and handmade mask/surgical mask. Individual statistical tests were performed for each distance from the source.

## 3. Results

### 3.1 IMAGING DROPLETS IN FLIGHT

We recorded images at eight different positions, whose vertical and horizontal distances are presented in Fig. 1A. Fluorescent droplets appeared as segments on the images, with a length proportional to their speed (Figure S2). We observed three types of droplets: droplets smaller than ~30 μm that remained airborne following air currents in the room, droplets larger than ~170 μm that fell ballistically with trajectories similar to the red lines in Figure 1A, and intermediate size droplets which could show any of the two behaviours or fall with a non-ballistic trajectory. The size of the droplets was assessed using shadow imaging technique (Supplementary Material).

Visually, the impact of placing a mask on the manikin was obvious, especially under coughing conditions (Figure 1B). To quantify the data, we counted the number of particles crossing the lower edge of the field of view at positions 1 to 7, therefore extrapolated to have deposited on the table. Data were compiled from 100 images taken from six replicate runs, each with no face covering, with a surgical mask or with a single-layer face covering. We used a new mask for each repeat to account for potential variability in the masks. The absolute particle count was divided by the length of the lower edge of the field of view and by the total period of observation to provide the rate of deposition of particles along the centreline of the table. Tests modelling speaking without a mask showed an exponential decline in particle deposition rate with distance, with none reaching the 2 m mark (Fig. 1C), while the distribution peaked at 1 m and dropped more slowly for coughing (Fig. 1D). When the manikin was fitted with either surgical or handmade masks, not one droplet was found to fall through the lower edge of the fields of view in either speaking or coughing simulations. Therefore, both face masks were largely impermeable to droplets larger than 20-30 μm.

### 3.2 DROPLET DEPOSITION

To provide an overview of the spread of droplets over the table, we first used UV light to image the distribution of fluorescent particles that accumulated on white paper placed in front of the manikin. An example result for a speaking test is shown in Figure S3. We measured droplet deposition rate according to distance, presented as the number of particles divided by the sampling area and the duration of the experiment by counting the droplets every 5 and 15 cm from the manikin for speaking and coughing, respectively. Each experiment was performed six times with and without both types of face-covering, using a new mask for each repeat. The numbers of surface-deposited droplets were consistent with the measurements of falling droplets (Figure 2A, B). Once again, the presence of a face-covering very effectively blocked droplet deposition. However, because we measured droplets on a larger area and longer duration (up to 20 min) than the flight tests, we could capture less frequent events. With both mask types, small numbers of droplets (up to seven) were seen at various distances from the manikin, but these values were four and five orders of magnitude lower than those observed without a mask for speaking and coughing, respectively. Hence, these represent events with a frequency of one in 10,000 and one in 100,000, respectively.

To further corroborate our results, we imaged deposited droplets using microscopy. To this end, the manikin was set to emulate speaking or coughing as before and fluid droplets were collected on glass slides positioned at varying distances (0.25, 0.5 and 0.75 m for speaking conditions and 0.5, 1, 1.5 and 2 m for coughing conditions) from the manikin. We repeated each experiment six times each with and without masks, using a new mask for each test. After each run the glass slides were collected and imaged with a 20X objective to count the number of droplets deposited in both cases. We then reconstructed the spatial distribution of droplets with and without masks (Figure 2 C, D). Consistent with other means of measurement, both types of face covering were extremely effective at reducing the particle deposition rate. As with UV imaging of droplets deposited on white paper, this technique also allowed a long experimental duration (20 min) and thus the capture of statistically rare events. With both mask types, we observed a few (up to 4) droplets at various distances from the manikin, including at 2 m for coughing. However, this number was again 4 - 5 orders of magnitude lower than the number of droplets observed without a mask for speaking and coughing, respectively.

### 3.3 DROPLET DEPOSITION FROM HUMAN VOLUNTEERS

Results obtained from a manikin, one might argue, are only indicative due to the wide variability in the expiration parameters for speech and coughing in humans [16,17]. Accordingly, we performed experiments using four male and two female volunteers (aged between 30 and 45) to either read a sample script for three minutes or cough for one minute. For each test, a glass slide was placed vertically, 5 cm in front of the individual’s mouth. At the end of each experiment, the sample was imaged using widefield microscopy to count the droplets generated by the subject. Each speaking and coughing test was performed twice each with and without a surgical mask on separate days, reversing the order of the tests on day two. In contrast to the manikin tests, there was a large variability in the number of droplets emitted by the individual subjects in the absence of a mask (Figure 3). However, for all subjects, we did not find a single droplet when a mask was worn.Speech

**Figure 1.**
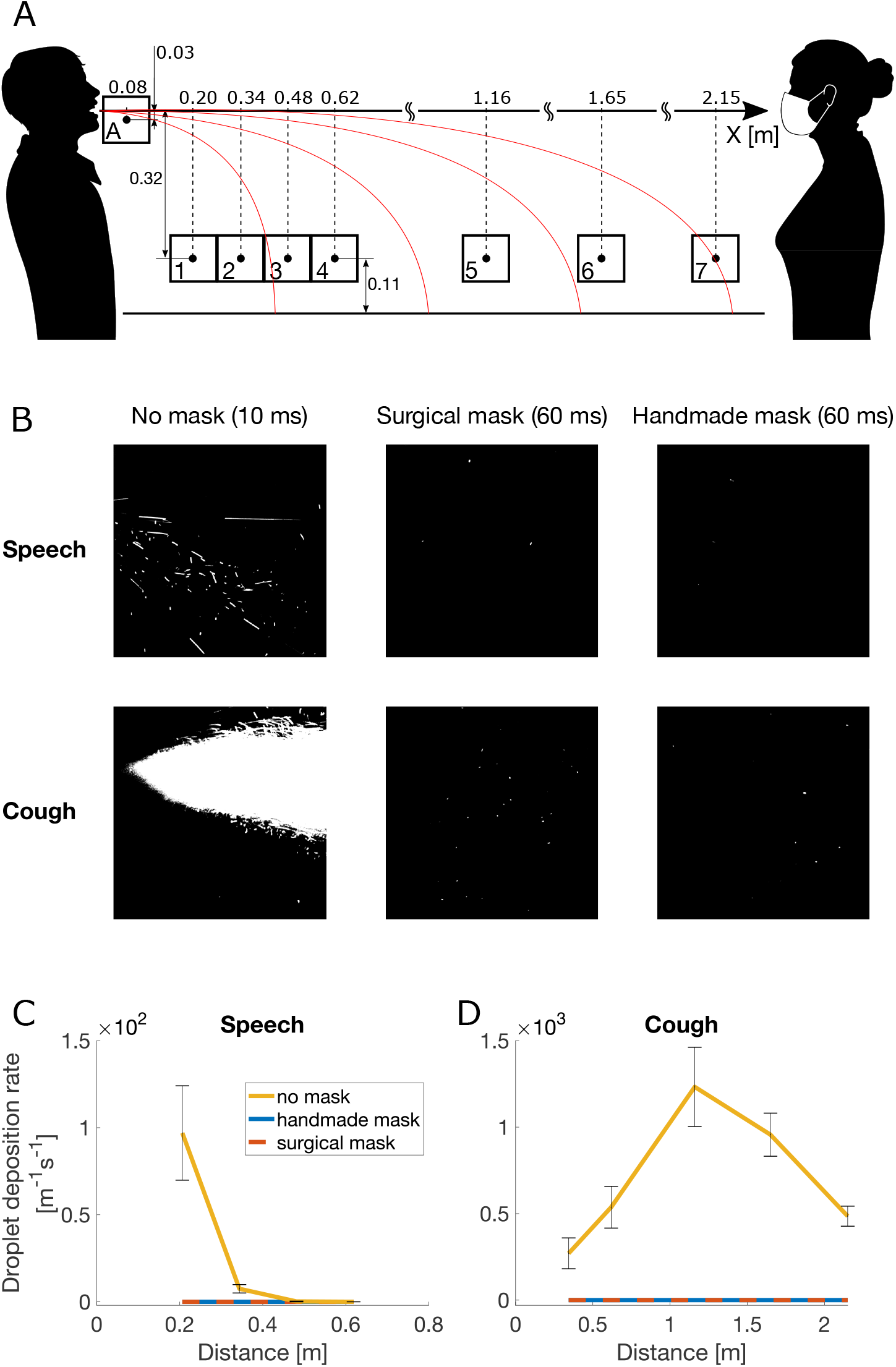
Laser Imaging of Respiratory Droplets in Flight. (A) Schematic diagram of the experimental setup. Boxes indicate imaging windows. (B) Examples of images captured at position A (directly in front of the mouth) for speaking (upper row) and coughing (lower row), without mask (1^st^ column), with the handmade mask (2^nd^ column) and with the surgical mask (3^rd^ column). (C, D) Droplet deposition rate over the table centreline versus the horizontal distance from the manikin’s mouth in (C) speaking and (D) coughing conditions. Data are the mean ±1 SEM of six independent replicates. (C) Both mask types statistically significantly reduced the droplet deposition rate at 0.21 m from the mouth of the source (p = 0.016 in both cases). Past 0.21 m, too few droplets were detected also without mask to observe statistically significant differences among the tested conditions. (D) No significant differences were detected between mask types, but both mask types statistically significantly reduced the droplet deposition rate at every tested position (p = 0.019).

**Figure 2.**
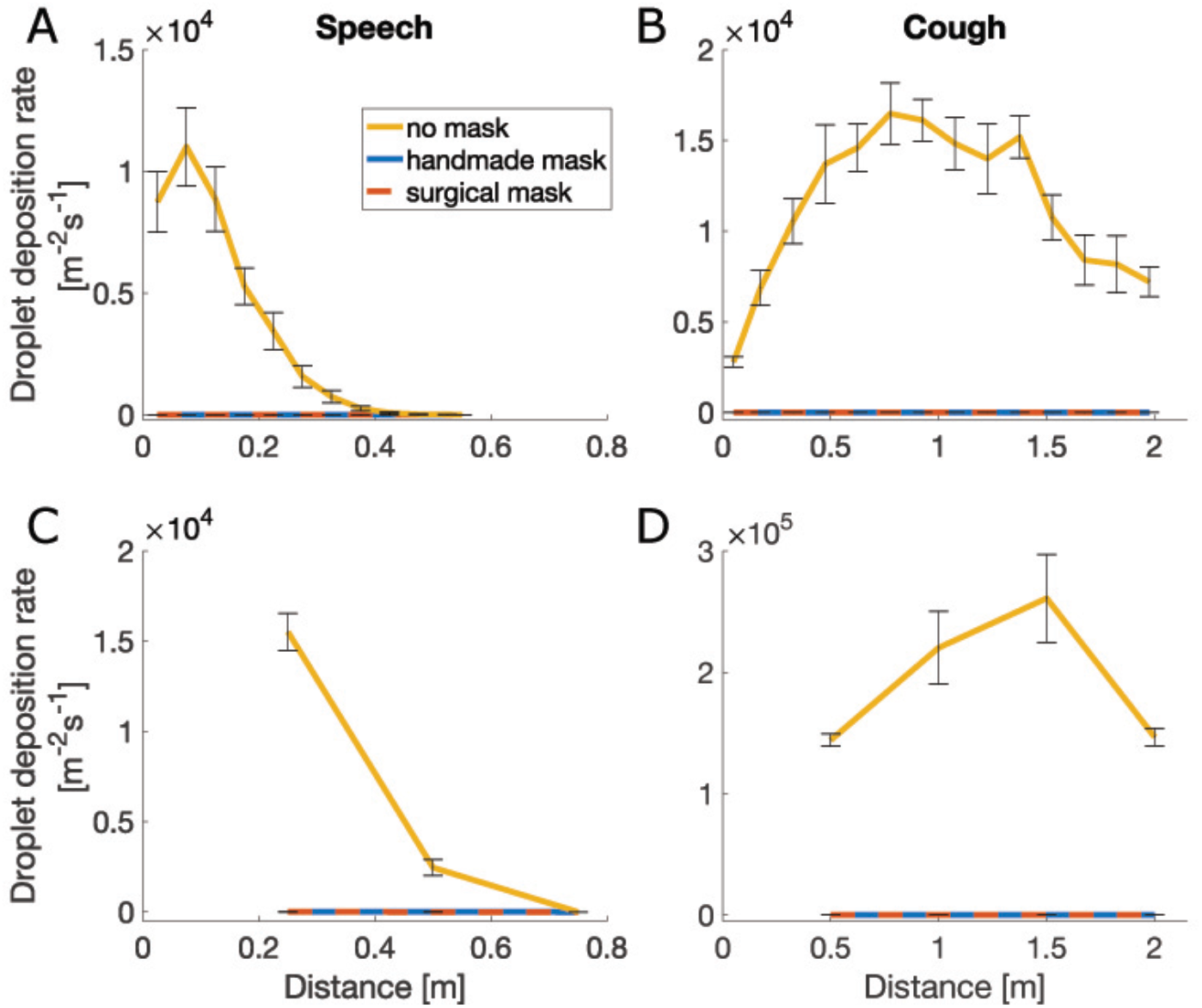
Surface Deposition of Droplets. Droplet deposition rate versus horizontal distance from the manikin’s mouth in (A, C) speaking or (B, D) coughing conditions, measured by (A, B) UV illumination of paper sheets or (C, D) microscopic imaging of glass slides placed on the table centre line. Data are the mean ± 1 SEM of six independent repeats. Statistical analyses indicated that for (A), both mask types significantly reduced the droplet deposition rate at up to 47.5 cm from the source (p = 0.047). For (B), droplet deposition rate with and without masks were not significantly different (p = 0.055). For (C), both mask types significantly reduced droplet deposition rate up to 0.5 m from the source (p = 0.012). Past that point, droplets were undetectable in all cases. For (D), both mask types significantly reduced droplet deposition rate at all distances from the source (p = 0.016). No statistically significant difference was observed between the two mask types in any test.

**Figure 3.**
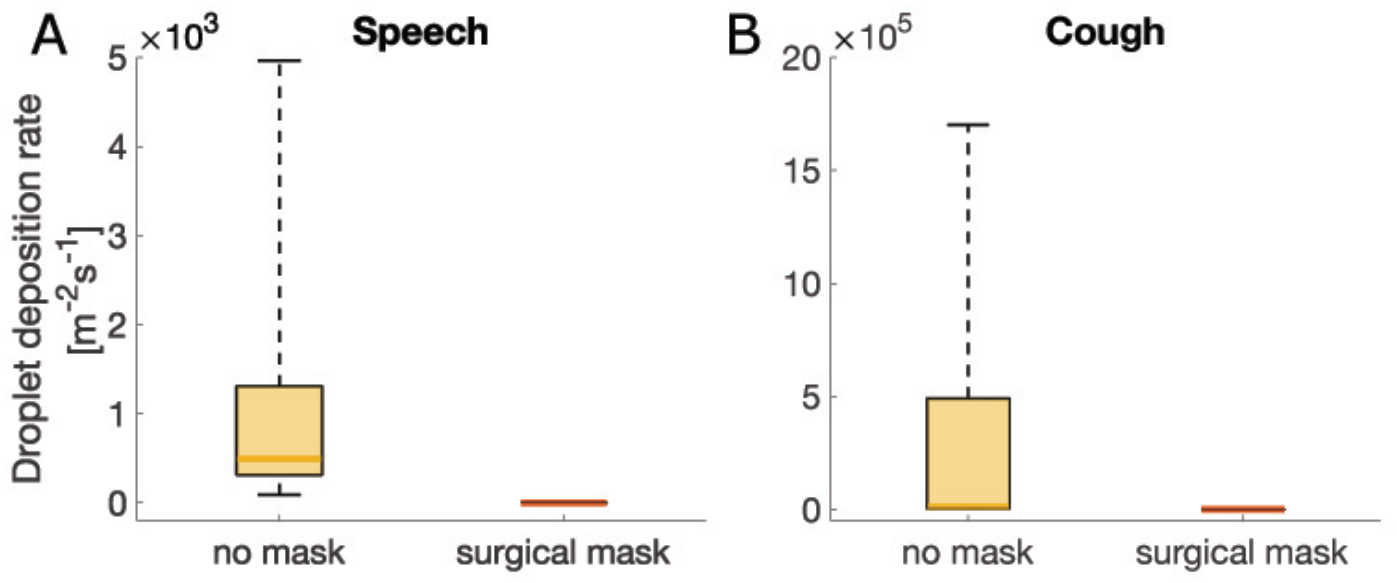
Droplet Deposition from Human Volunteers. Droplet deposition rate in human volunteers for the no-mask and surgical mask cases under (A) speech and (B) coughing conditions. Measurements are plotted as a box between the 25^th^ and 75^th^ percentiles with a mark indicating the median. The whiskers extend over the full data range. Statistical tests showed p = 2.3 × 10^−6^ for both cases, thus rejecting the null hypothesis that the two samples were drawn from the same distribution. Therefore, the use of surgical masks statistically significantly reduced droplet deposition rate.

## 4. Discussion

We employed three independent, quantitative techniques to measure the droplet deposition rate at increasing distance from a manikin. Without any face covering, we measured distributions of deposition rates comparable to those reported in the literature [18-20]. When the manikin wore any of the two face masks, we observed that less than one in 1,000 particles escaped for both speaking and coughing. To further demonstrate that our results could be applied to real people speaking and coughing, we tested six individuals using microscopy. Without masks, we measured between 10s and 1000s of particles for speaking and coughing without a mask, respectively. Conversely, we found zero particles with the surgical mask, both for speaking and coughing.

Overall, these data demonstrate that face masks are highly effective at reducing exhalation of large respiratory droplets. As these droplets are likely to be the main driver of SARS-CoV-2 transmission [2,3,8], our data suggest that the wearing of masks can substantially reduce the probability of an infected person transmitting the virus. In this study, however, we focused only on the large respiratory droplets that land on a surface within few seconds. Other studies such as, for instance Fisher et al. [13], which focuses on in-flight droplets and that include smaller droplets and aerosol, are likely to show a lower level of effectiveness depending on the fabric type. Hence, if aerosol transmission is later determined to be a significant driver of infection [21], then our findings may overestimate the effectiveness of face coverings.

Air was ejected at velocities and flow rates within the range of those observed from real individuals [14,15] but droplets were ejected at a higher volume rate. Hence, the physical interaction between the droplets and the airflow jet [16,20] was not correctly represented. However, this was necessary to ensure a high volume of particles and thus robust statistics, in a sufficiently short time frame to minimise evaporation of landed droplets and contamination with other particles in the environment. By counting from 1 to 100, a person ejects between 10 and 100 mg of water [19], whilst our manikin ejected about 10 mg s^−1^. Thus, each second of test corresponded to a person counting at least from 1 to 100. The duration of the tests was as high as 20 minutes; hence we modelled a person counting up from 1 to more than 10,000. In one cough a person ejects about 1 mg of fluid [19], whilst our manikin ejected 100 mg s^−1^. We tested cough for as long as 10 minutes, which is equivalent to 60,000 coughs. Furthermore, it was not possible to test if droplets escaped the mask and landed outside of the table. Viola et al. [10] showed that surgical and handmade masks can lead to lateral and backwards aerosol jets. If a large droplet was carried by these jets, we would have not detected it. Finally, we tested simple single layer handmade masks, but it should be recognised that there is a wide range of handmade masks and some might not be fit for purpose.

In conclusion, these experiments demonstrate that both surgical and simple handmade masks such as a single layer cotton mask are can suppress the risk of direct person-to-person virus transmission through large droplet deposition. The data do not allow us to draw conclusions on the risks of virus transmission through aerosol inhalation. Assuming that SARS-CoV-2 virus transmission through aerosol is small compared to through large droplets, these results suggest that physical distancing can be reduced with the use of face coverings.

## Data Availability

All metadata and raw data are available on Edinburgh DataShare

https://datashare.is.ed.ac.uk/

## Acknowledgements

The authors are grateful to Dr B. Peterson (School of Engineering) and Dr A. Nila (LaVision) for their advice on the laser tests and for lending related equipment, and to Dr K. Dunn (School of Engineering), Prof. F. Denison (Queen’s Medical Research Institute) and Dr F. Mehendale (Usher Institute) for procuring the tested face masks.

Bandiera is supported by the UK Engineering and Physical Sciences Research Council (EPSRC), grant no. EP/P017134/1. Pavar and Pisetta are supported by the EPSRC grant EP/L016680/1, while Molinari by the EPSRC grant EP/S02431X/1. Ōtomo’s scholarship is funded by the Japan Student Services Organization. Menolascina is supported by the European Commission (766840) and the EPSRC (EP/S001921/1 and EP/R035350/1), whilst Digard is funded by the UK Biotechnology and Biological Sciences Research Council (BB/P013740/1).

## Author Contributions

Bandiera, Menolascina, Otomo, Pavar and Pisetta undertook the experiments. Mangano led the design and manufacturing of the speech and cough simulator. Menolascina conceived and led the microscopy investigation. Digard, Menolascina, Molinari, Seckl and Viola designed the experiments. Viola coordinated the project. Molinari wrote the first draft of this document that was edited, reviewed and approved by all the authors.

## Supplementary Information

TABLE OF CONTENTS

S1. MASKS TYPES AND MATERIALS

S2. LASER-IMAGING OF DROPLETS IN FLIGHT

S3. DROPLET DEPOSITION

S4. CHARACTERISING PARTICLE SIZE AND VELOCITY

### S1. MASKS TYPES AND MATERIALS

The surgical masks were “Disposable Medical Masks” purchased through the University of Edinburgh’s Pharmacy from Henan Yunda Medical Equipment Co Ltd., 158 Dinzhang Road, Changyuan City, Xinxiang City, Henan Province, China (Certificate of Compliance to EN 148:2001 and A1:2009 Standard, number 0P200315.HYMUT56). The handmade masks were “Reusable Fabric Face Mask, White, One Size” purchased through Amazon (item model number: MSK10UK; ASIN: B08888C81Z) from FM London Accessories, Unit 13, Basset Court, Loake Close, Grange Park, NN45EZ, UK.

To gain further insights into the masks we used in our tests, we sought to obtain micrographs of their plots. To this aim we excised small sections of the masks (roughly 1 cm in width and 2 cm in length) and attached them to a microscopy glass slide to proceed to imaging via phase-contrast micrography. We used a CFI Plan Fluor 4X objective mounted on a Nikon Eclipse Ti inverted microscope, equipped with an Andor Zyla sCMOS camera, resulting in a resolution upon magnification of 1.63 μm/pixel. A field of view of 2.77 mm^2^ in total, was acquired in bright-field using NIS-Elements imaging software. The interwoven cotton threads of hand-made masks (Figure S1A) leave gaps of around 50 μm (white spots). Surgical masks are formed from multiple layers of fibres with a broader spectrum of gap sizes (Figure S1B).

**Figure S1.**
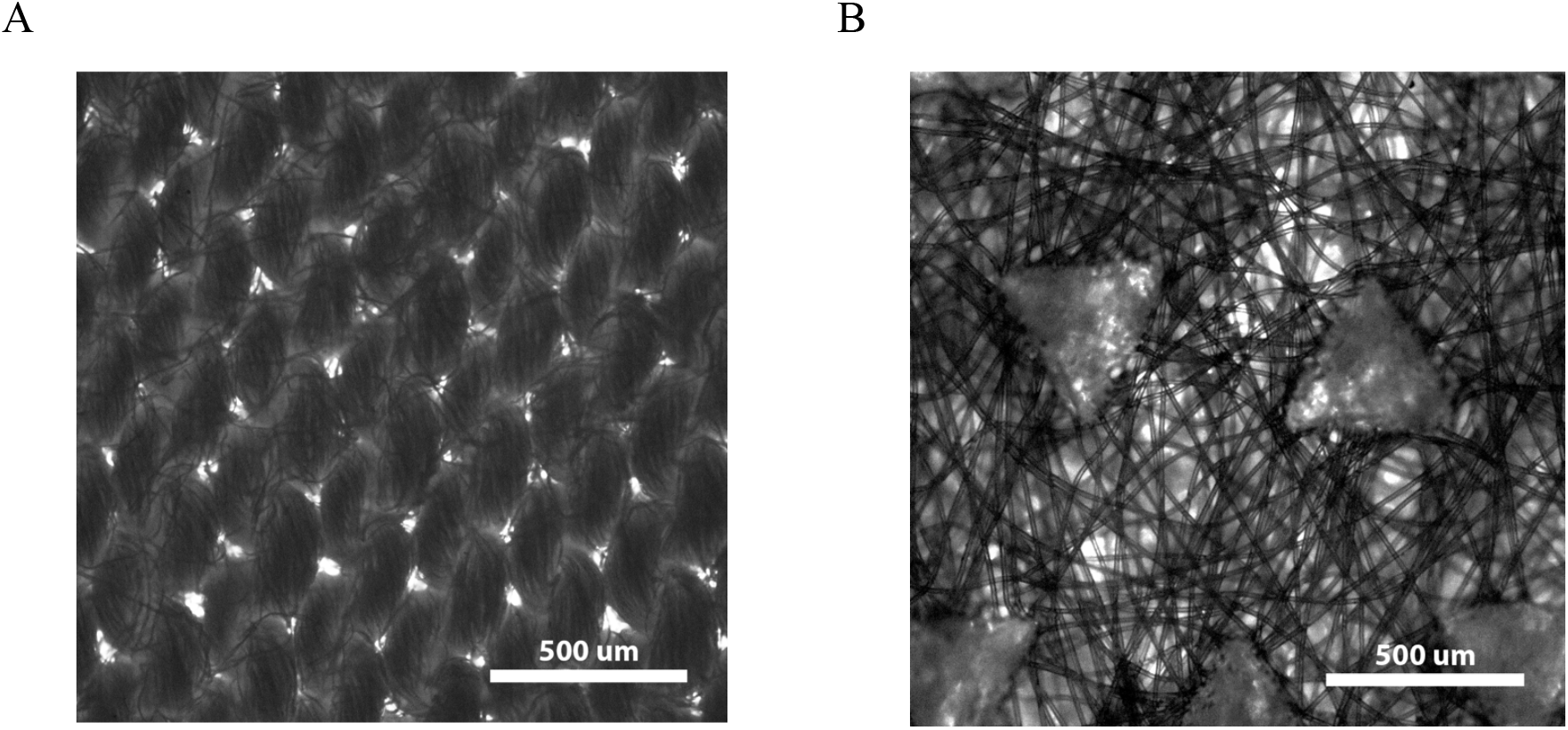
Micrographs of Handmade (A) and Surgical Masks (B). The structural properties, and differences, of the plots are clearly visible in these images: brighter spots highlight “pores”, i.e. points where the individual would be exposed to suitably small particles from the outside.

To image larger respiratory droplets in flight, we projected a thin laser sheet along the vertical plane through the mouth of the manikin and used a photographic camera to capture the light scattered by droplets passing within this plane (Supplementary Movie 1).

To capture images for analysis, the camera was placed at varying positions from the manikin. Position A (Figure 1A) was located directly in front of the mouth, with its centre at (x, y) = (0.085, -0.027) m, relative to the centre of the mouth. The centre of the other positions (1-7) were at y = -0.32 mm in the vertical direction. The centre of positions 1, 2, 3, 4, 5, 6, and 7 were at x = 0.2 m, 0.34 m, 0.48 m, 0.32 m, 1.16 m, 1.65 m, and 2.15 m in the longitudinal direction, respectively. Positions 1-4 were used for speaking conditions, and positions 2, 4, 5-7 were used for coughing conditions. Fluorescent droplets appeared as segments on the images, with a length proportional to their speed (Figure S2).

**Figure S2.**
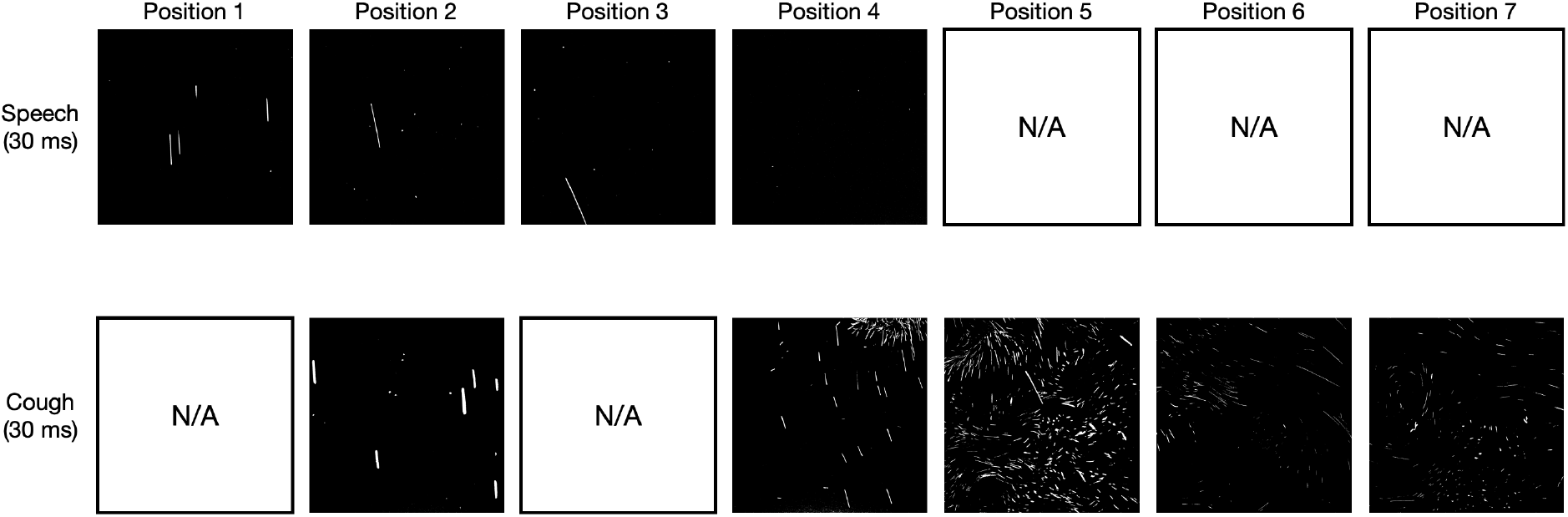
Examples of In-Flight Droplets. Examples of images captured at positions 1-7 (directly above the table) for speaking (upper row) and coughing (lower row).

### S3. DROPLET DEPOSITION

To provide an overview of all of the droplets deposited from the manikin, we used UV light to image the distribution of fluorescent particles that accumulated on white paper placed in front of it. An example result is shown in Figure S3A, which shows the droplet distribution observed after a speaking test. We also used microscopy to corroborate the results obtained with UV light imaging. A sample image, acquired on a slide placed 25 cm from the manikin, is shown in Figure S3B.

**Figure S3.**
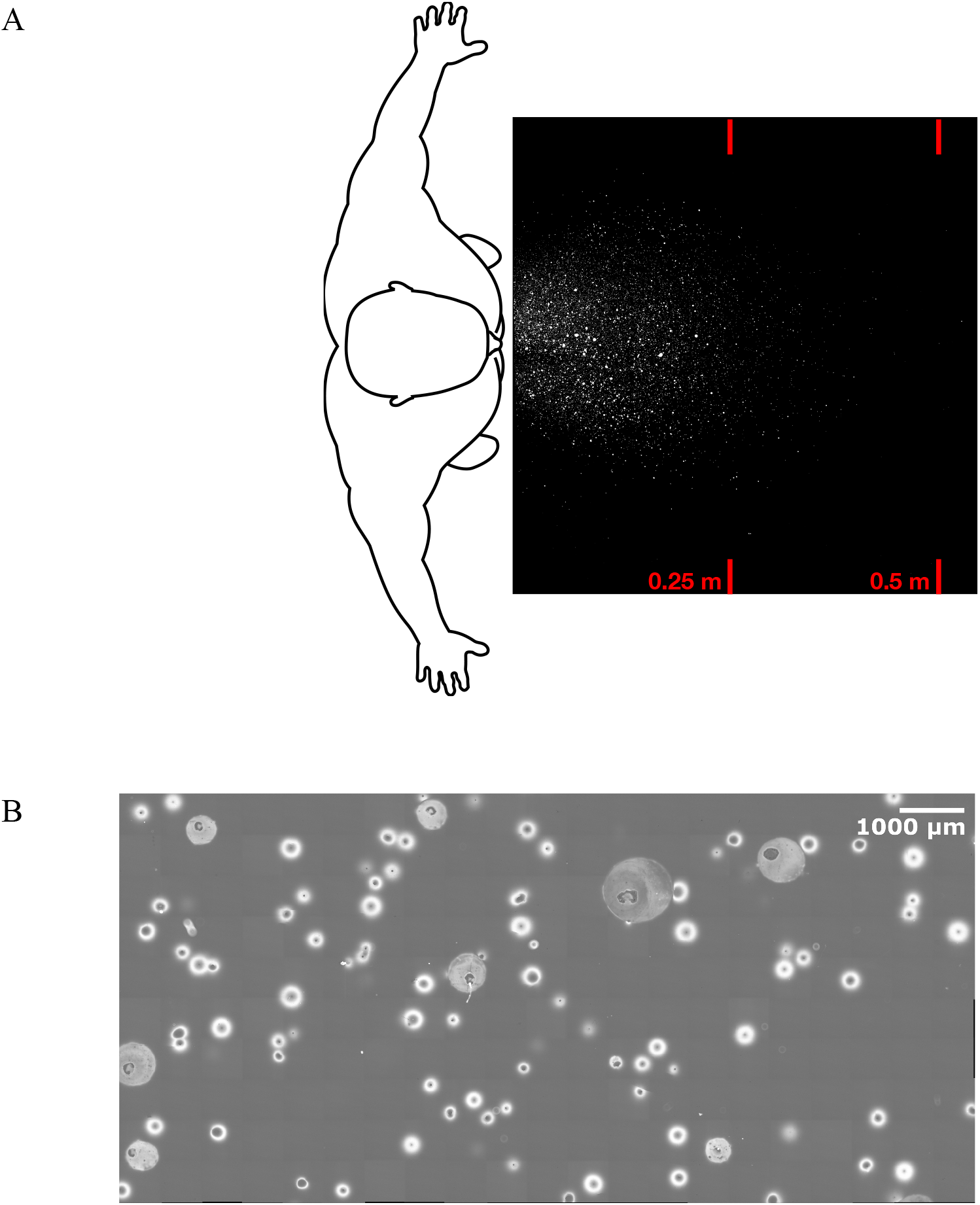
Example of Droplet Distributions on the Table. Droplets found on the table for a speaking test in absence of face covering measured Using UV light imaging (A) and microscopy (B).

### S4. CHARACTERISING PARTICLE SIZE AND VELOCITY

We used shadow imaging to characterise particle size range and behaviour. An 8-bit CCD camera with a resolution of 2056 × 2060 pixels with a Tamron 180 mm F3.5 SP AF Di Macro Lens was used to image a physical plane of 26.96 × 27.01 mm. This gives a resolution of 13.1 μm/pixel, so the following particle diameters are given to ± 7 micron. A fluorescent light was used as a backlight and a light diffuser was used to create a uniform background (Figure S4). Water droplets coloured by black food dye were visualised. Measurements were taken at seven positions: A, B, 2’, 4’, 5’, 6’, and 7’. Position A was located directly in front of the mouth, as for the laser tests, with its centre at (x, y) = (0.085, -0.027) m, relative to the centre of the mouth (Figure 1A). Position B was 0.2 m above position A at (x, y) = (0.085, 0.173) m. The centre of positions 2’, 4’, 5’, 6’, and 7’ were located 45 mm below the centres of the corresponding laser test positions 2, 4, 5, 6 and 7, respectively (y = -0.365 m). Images were recorded with apertures f/3.5, f/8, f/11 and f/16 combined with exposure times of 0.07, 0.5, 1 - 1.5 and 2 ms, respectively.

**Figure S4.**
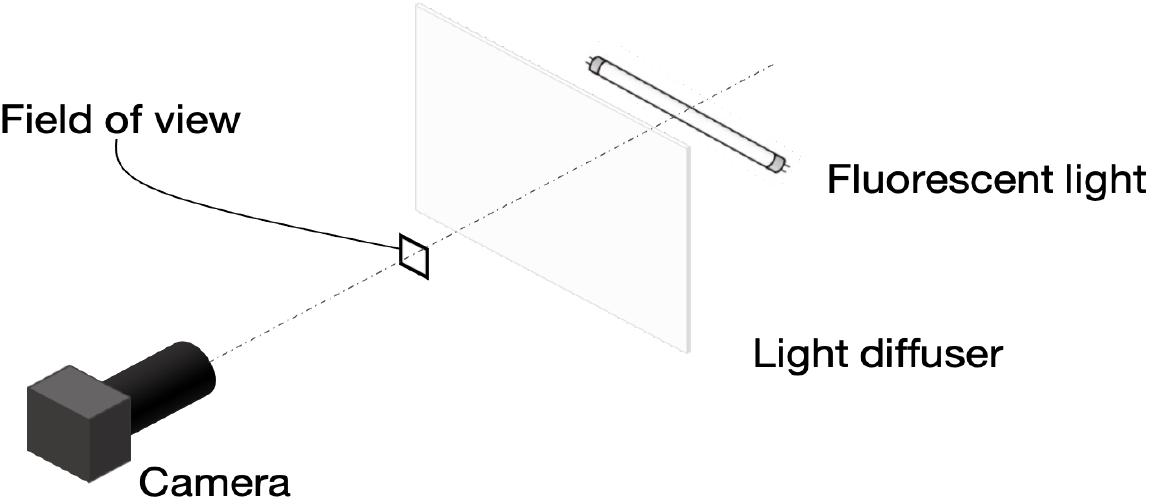
Experimental Setup for Shadow Imaging.

The black coloured water droplets appeared as segments with thickness equal to their diameter and length proportional to their speed (Figure S5). Overall, the smallest and largest particles seen were 26 ± 7 μm and 590 ± 7 μm, respectively, and the largest particles displaying non-ballistic behaviour were 170.3 ± 7 μm.

Particle behaviour was affected by the background flow to an extent depending on both particle size (smaller particles being more affected) and the strength of the flow. With shadow imaging, we could differentiate between particles behaving ballistically or non-ballistically based on pathline angle (Table S5). Specifically, measured anti-clockwise from the positive x-axis (Figure 1), pathline angles smaller than 0 and less than or equal to -90 degrees were considered to be ballistic, while angles greater than or equal to zero and less than 90 degrees were considered non-ballistic. From the laser imaging using a wider field of view, we could both visualise flow eddies and track some particles between frames, which enabled us to distinguish floating and falling behaviours.

From the above results, we draw the following conclusions. First, we can see particles smaller than 26 ± 7 μm with the laser. Second, we can measure the size of droplets as small as 26 ± 7 μm with shadow imaging and see that these often follow the background flow instead of falling. Third, we can see that droplets up to 170 ± 7 μm sometimes travel non-ballistically and that the larger droplets are only minimally deviated from a ballistic trajectory. Hence, for the laser particle tests, we concluded that we counted all droplets that fell ballistically and that we had a resolution that allowed us to see droplets small enough that they did not fall but followed air currents instead.

**Figure S5.**
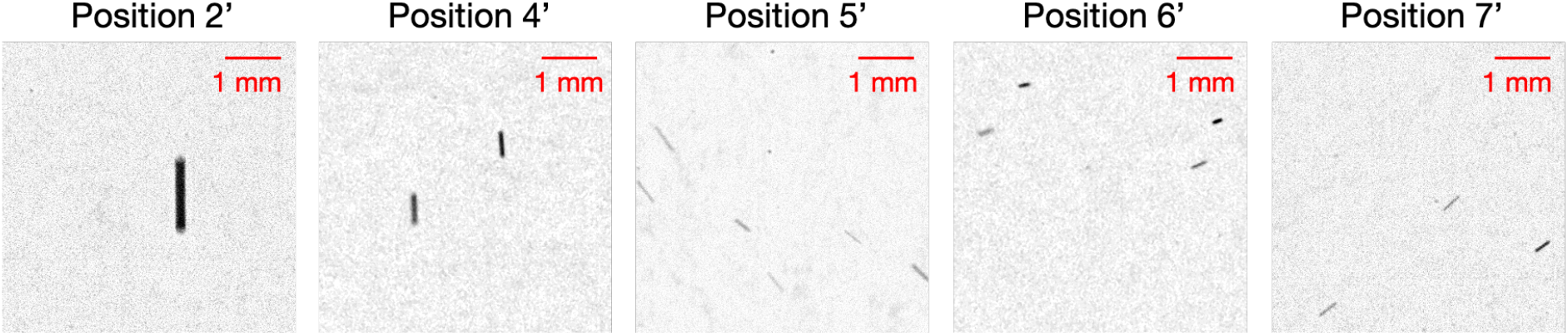
Examples of Shadow Images. Example of images captured at positions 2’, 4’, 5’, 6’, and 7’ using an exposure time of 2 ms. The images are cropped with 5.24 mm × 5.24 mm.

**Table S5:** Particle Behaviours and Diameters. All values are ± 7 μm.

| Position | Particle size ($\mu\text{m}$) | | |
| --- | --- | --- | --- |
|  | Overall | Non-ballistic | Ballistic |
| B | < 26 | < 26 | < 26 |
| A | 26 - 590 | 26 - 144 | 26 - 590 |
| 2 | 39 - 550 | 52 - 170 | 39 - 550 |
| 4 | 26 - 301 | 26 - 144 | 39 - 301 |
| 5 | 26 - 341 | 26 - 92 | 26 - 341 |
| 6 | 26 - 223 | 26 - 144 | 26 - 183 |
| 7 | 26 - 131 | 26 - 52 | 26 - 131 |

